# Two independent introductions of SARS-CoV-2 into the Iranian outbreak

**DOI:** 10.1101/2020.11.16.20229047

**Authors:** Zohreh Fattahi, Marzieh Mohseni, Khadijeh Jalalvand, Fatemeh Aghakhani Moghadam, Azam Ghaziasadi, Fatemeh Keshavarzi, Jila Yavarian, Ali Jafarpour, Seyedeh elham Mortazavi, Fatemeh Ghodratpour, Hanieh Behravan, Mohammad Khazeni, Seyed Amir Momeni, Issa Jahanzad, Abdolvahab Moradi, Alijan Tabarraei, Sadegh Ali Azimi, Ebrahim Kord, Seyed Mohammad Hashemi-Shahri, Azarakhsh Azaran, Farid Yousefi, Zakiye Mokhames, Alireza Soleimani, Shokouh Ghafari, Masood Ziaee, Shahram Habibzadeh, Farhad Jeddi, Azar Hadadi, Alireza Abdollahi, Gholam Abbas Kaydani, Saber Soltani, Talat Mokhtari-Azad, Reza Najafipour, Reza Malekzadeh, Kimia Kahrizi, Seyed Mohammad Jazayeri, Hossein Najmabadi

## Abstract

The SARS-CoV-2 virus has been rapidly spreading globally since December 2019, triggering a pandemic, soon after its emergence, with now more than one million deaths around the world. While Iran was among the first countries confronted with rapid spread of virus in February, no real-time SARS-CoV-2 whole-genome tracking is performed in the country.

To address this issue, we provided 50 whole-genome sequences of viral isolates ascertained from different geographical locations in Iran during March-July 2020. The corresponding analysis on origins, transmission dynamics and genetic diversity, represented at least two introductions of the virus into the country, constructing two major clusters defined as B.4 and B.1*. The first entry of the virus occurred around 26 December 2019, as suggested by the time to the most recent common ancestor, followed by a rapid community transmission, led to dominancy of B.4 lineage in early epidemic till the end of June. Gradually, reduction in dominancy of B.4 occurred possibly as a result of other entries of the virus, followed by surge of B.1.* lineages, as of mid-May.

Remarkably, variation tracking of the virus indicated the increase in frequency of D614G mutation, along with B.1* lineages, which showed continuity till October 2020.

According to possible role of D614G in increased infectivity and transmission of the virus, and considering the current high prevalence of the disease, dominancy of this lineage may push the country into a critical health situation. Therefore, current data warns for considering stronger prohibition strategies preventing the incidence of larger crisis in future.

## Introduction

The coronavirus disease 2019 (COVID-19) pandemic, caused by severe acute respiratory syndrome coronavirus 2 (SARS-CoV-2) has now crossed one million deaths while infected 51,259,771 people worldwide as of 10 November 2020. The SARS-Cov-2 outbreak first emerged in a seafood and animal market in Wuhan, Hubei Province, China, in late December 2019 [1]. The first reported case out of China was detected in Thailand on January 13^th^ 2020 and the pandemic was announced by The World Health Organization (WHO) on March 11^th^ 2020.

Real-time whole-genome sequencing (WGS) of emerging viruses can provide valuable information about the transmission dynamics of the epidemics and, if combined with epidemiological data, can accurately evaluate the magnitude of transmission and therefore offer insights for management and efficient responses to control the outbreak in a region [2]. Along with this knowledge, real-time whole-genome sequencing of this emerging virus was commenced at the early phase of outbreak globally and an increasing number of sequences are being deposited in the global initiative on sharing all influenza data (GISAID) [3]. In addition to the outbreak tracing and investigating the phylodynamics and evolutionary events of SARS-CoV-2, these data provide beneficial information for adapting more specific diagnostic tests, therapeutic approaches and vaccines.

Iran was among the first countries confronting the rapid virus spread, which in a little while became a concerning issue in the country. The first COVID-19 confirmed patient and death report from the disease was announced on a same day on 19 February 2020 in Qom city. Local transmission to the other neighboring provinces was reported just a day after and then the disease spread shortly Iran-wide. The first outbreak peak dropped in April, but soon after relaxing the lockdown, the country experienced another increase in the number of cases in May. As of 10 November, 692,949 infected cases and 38,749 deaths have been reported officially and there is a concern about the increase in mortality rate in autumn, which is now estimated to around ∼7% according to official statistics (https://www.worldometers.info/coronavirus/country/iran/). Although the outbreak initiation in Iran was among the first ones in February along with the Italy and South Korea outbreaks, no real-time SARS-CoV-2 whole-genome tracking has been performed in the country so far. The first whole-genome sequence of the virus from Iran was deposited in GISAID database (EPI_ISL_424349) on 4 April 2020. In the same month, the study by Eden et al. revealed three major substitutions of G1397A, T28688C, and G29742T in genomes of patients with travel history to Iran, which constitute a distinct clade representative of the specific viral diversity present in Iran at that time [4].

To date, there are only eight complete genomes available in GISAID, not sufficient for tracking the virus in the country. Therefore, we performed genome-sequencing of 50 SARS-CoV-2 samples ascertained from different geographical locations and time intervals in Iran. We aimed at extending the understanding of the origins and transmission dynamics, circulating lineages and variation tracking of SARS-CoV-2 outbreak in Iran, using molecular and phylogenetic methods.

## Material and Methods

### Specimen Recruitment

We recruited 50 SARS-CoV-2 RNA samples, obtained as part of clinical testing in different COVID-19 referral centers of the following provinces in Iran; Alborz (n=2), Ardabil (n=4), Gilan (n=6), Tehran (n=27), Khuzestan (n=3), Qom (n=1), Sistan and Baluchestan (n=3) and South Khorasan (n=4). All patients had been referred between March-July 2020 with the clinical presentations of the COVID-19 disease and were confirmed as SARS-CoV-2 positive by real-time RT-PCR assay at the corresponding local centers.

### Sequencing and Genome assembly

Whole-genome sequencing of SARS-CoV-2 RNA samples was performed by targeted enrichment including reverse transcription (RT) reaction for cDNA synthesis, followed by target enrichment and library preparation using CleanPlex® SARS-CoV-2 Research and Surveillance Panel (Paragon Genomics, Inc.). All samples were paired-end sequenced on an Illumina MiSeq instrument using 300-cycle MiSeq v2 reagent kits (Illumina, Inc.) and eventually 5.4 Gb of raw data with 94.5% of bases with >=Q30 were generated. Initially, the quality of FastQ files was assessed using the FastQC program [5] and then preprocessed using Fastp [6]. The sequences were aligned to the SARS-CoV-2 reference genome (NC_045512.2) using Bowtie2 [7], keeping the reads mapped in proper pair. The resultant filtered bam files were used for assembly of consensus SARS-CoV-2 sequences using samtools mpileup and bcftools[8]. Finally, the consensus FastQ files were converted into Fasta format using the seqtk program (https://github.com/lh3/seqtk), masking bases with quality lower than 20 to ambiguous nucleotides (N).

### Lineage assignment

In addition to the 50 sequenced samples in this project, 8 other full genome SARS-CoV-2 sequences from Iran in GISAID database, which were collected between March-May 2020, were subjected to lineage assignment. We tried Pangolin v2.0.7 (https://pangolin.docs.cog-uk.io/) [9], CoV-GLUE (http://cov-glue.cvr.gla.ac.uk/#home) [10] and NextClade v.0.6.0 (https://clades.nextstrain.org/) [11] to assign the global lineages present in Iranian SARS-CoV-2 outbreak. Pangolin and CoV-GLUE estimate lineages based on the methodology described by Rambaut et al. [9] as a dynamic nomenclature system to identify SARS-CoV-2 lineages. However, the NextClade estimates according to the five major clades used on Nextstrain (https://nextstrain.org/) presenting large scale and more stable diversity patterns. Therefore, we assigned both lineage nomenclature systems on our samples [12].

### Phylogenetic analysis

We used BEAST v1.10.4 for phylogenetic analysis and estimating the most recent common ancestor (TMRCA) [13]. First, the consensus sequences of each sample were evaluated by NextClade v.0.6.0 [11] and the sequences containing >5% ambiguous nucleotides (N) and bearing private mutations more than the expected number, were excluded from the downstream analysis. To explore the temporal signal of the data, the remaining high quality Fasta files were aligned by MAFFT v7.407 using the FFT-NS-2 algorithm [14] and then a maximum-likelihood phylogenetic tree was built by applying IQ-TREE v2.1.1 with the GTR+gamma model [15]. The temporal signal was explored by TempEst providing a root-to-tip regression plot [16].

Eventually, BEAST v1.10.4 was applied to estimate TMRCA and construct a Bayesian phylogenetic tree of the remaining 45 sequences, plus Wuhan-1 patient (EPI_ISL_402125) as an outgroup, using a simple model consisting of HKYγ codon partitioned 1 + 2, 3 substitution model, strict clock and coalescent exponential growth tree prior. Maximum clade credibility (MCC) tree was then made using TreeAnnotator with 10% burn-in from two separate Markov chain-Monte Carlo runs, explored by Tracer v1.7.1[17] and combined by LogCombiner from the BEAST package [13].

To trace possible sources of SARS-CoV-2 entry into Iran, an additional phylogenetic tree was constructed based on a total of 261 samples including a list of high-quality genomes in GISAID [18] from the start of epidemic till the end of February, and random subsets of samples in GISAID during March-June interval available as of September 27 and selected using mothur [19].

### Variant analysis of SARS-CoV-2 genomes of Iranian outbreak

Variant analysis was performed by CoV-GLUE, relative to the reference sequence (NC_045512.1)[10]. In total, 53 samples were investigated after excluding the sequences bearing private mutations above the threshold. Moreover, to track the renowned D614G mutation frequency, Sanger sequencing of additional 67 SARS-CoV2 positive samples collected during July-October, was performed using the following primer pairs designed by ARTIC network [20]; 5’-CCAGCAACTGTTTGTGGACCTA-3’ and 5’-CAGCCCCTATTAAACAGCCTGC-3’.

## Results

### Genome assembly and Data availability

In this study, we obtained 44 high-quality SARS-CoV-2 sequences (>98% of the genome is complete) from the Iranian outbreak. The remaining 6 samples covered >82% of the NC_045512.2 genome sequence. These sequences have been deposited in the GISAID database (EpiCoV). The metadata information including the geographical location, collection date, the percentage of ambiguous nucleotides (N) for each genome, the percentage of genome coverage compared to NC_045512.2 and the exact lineages defined for each sample are provided in TableS1.

### Lineage assignment

Lineage assignment with Pangolin v2.0.7 and CoV-GLUE, although slightly different for some samples, both yielded B.4 as the dominant lineage circulating in Iranian SARS-CoV-2 outbreak till June; comprising 63.8% and 74% of all 58 samples studied through the March-July interval, respectively (see Figure 1A for spectrum of circulating lineages and TableS1 for exact lineage of each sample).

**Figure1.**
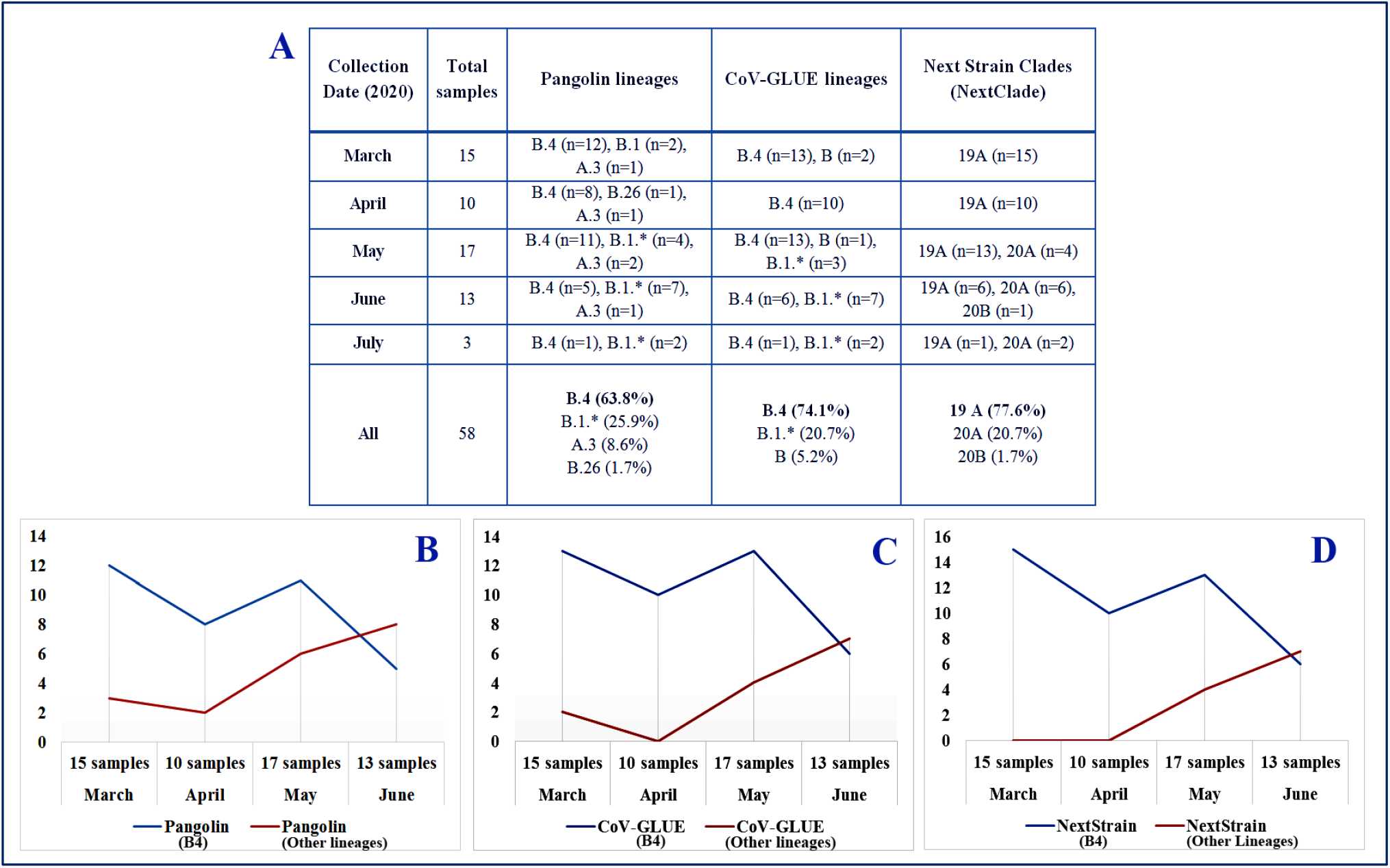
**A**. Lineages assignment of 58 SARS-CoV-2 sequences from the Iranian outbreak. Abundance of SARS-CoV-2 lineages over time from March to the end of June 2020 indicates a reduction in dominancy of the B.4 lineage. Lineage assignment by **B**. Pangolin v2.0.7, **C**. CoV-GLUE and **D**. NextClade v0.6.0

This is consistent with previous reports[4] as also the majority (83%) of SARS-CoV-2 sequences in GISAID that were primarily exposed in Iran, belonged to the B.4 lineage (TableS2).

In addition, the allocated lineages provided by NextClade v0.6.0 showed a dominancy of the 19A major clade (77.6%) which with 19B comprised the most prevalent clades during the early phase of the outbreak, especially in Asia. The dominancy of this clade in the Iranian outbreak is consistent with Iran being one of the first countries infected by the virus.

The A.3 lineages (Figure 1A) allocated by Pangolin are assumed to be not correct due to the lower quality of corresponding sequences. The B.4 lineage is more probable, as predicted by CoV-GLU and also supported by those samples all carrying the G1397A, T28688C, and G29742T substitutions [4].

Therefore, our results confirm B.4 as the dominant lineage in Iranian outbreak during the February-June interval and introduces B, B.1.* and B.4 as the current circulating lineages in the country.

On the other hand, sequencing more SARS-CoV-2 samples from the end of June onward is required to evaluate whether B.4 still persists as the dominant lineage. The current data (Figure 1B, 1C, 1D) already exhibit a trend towards the appearance of other SARS-CoV-2 lineages and reduction in dominancy of B.4. As of May, the 20A clade, being the dominant European clade in early 2020, started to appear and more B.1* lineages observed in the Iranian epidemic. This is explicable by new sources of virus entries to the country at that time interval. Its appearance could also be due to SARS-CoV-2 genome mutations in the existing Iranian B lineages, which were circulating alongside the B.4 lineage in early phase of the epidemic, although with a lower proportion.

### Phylogenetic Analysis

#### SARS-CoV-2 entry and circulating lineages

The phylogenetic tree of SARS-CoV-2 genomes from the Iranian outbreak clearly revealed two different circulating clusters (Figure 2), suggesting at least two separate introductions into the country.

**Figure2.**
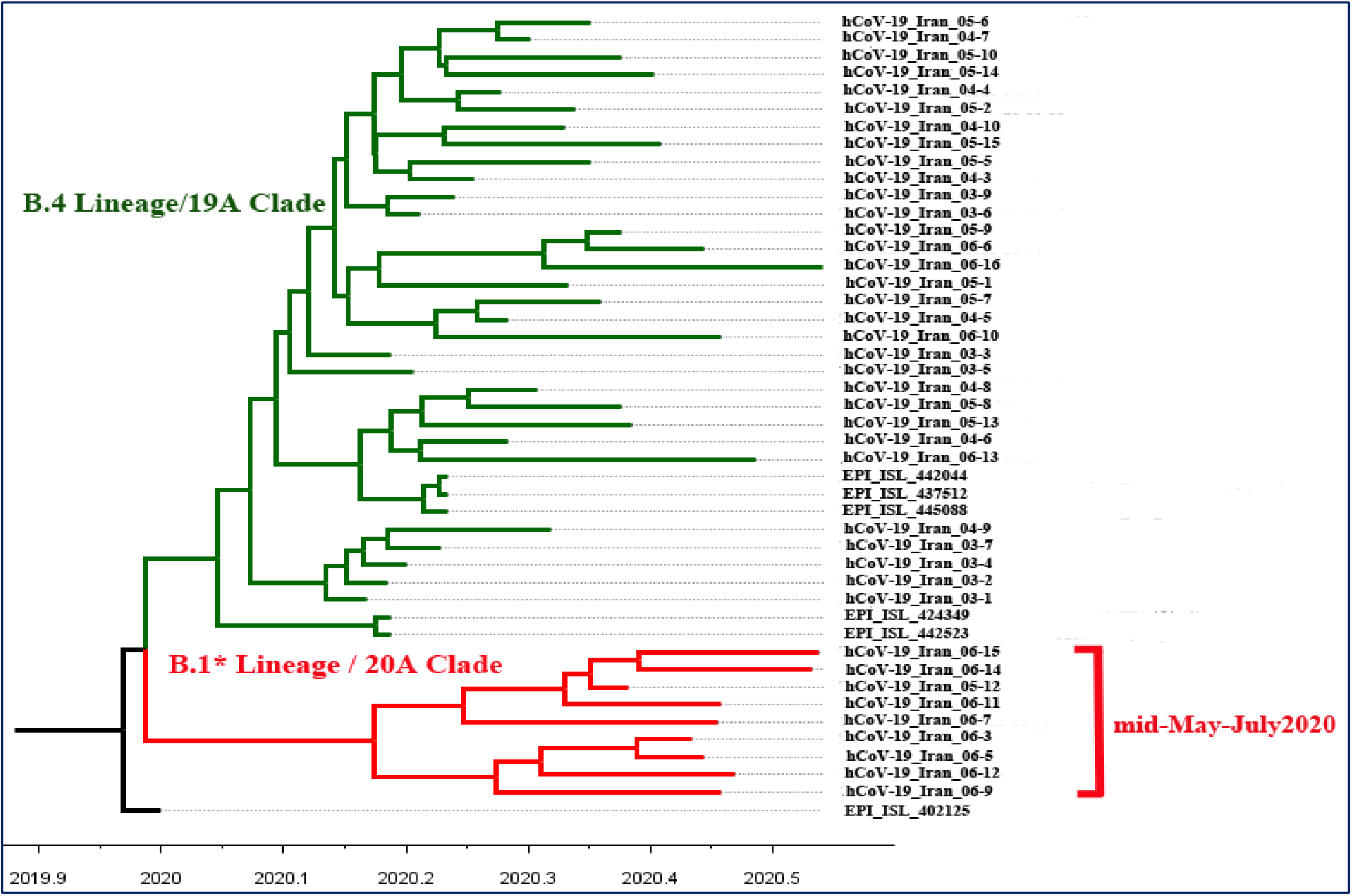
Tempo-spatial phylogenetic tree of SARS-CoV-2 emergence in Iran

The older green cluster is comprised of 36 genomes almost all of the B.4 lineage or 19A clade [B.4 / 19A], carrying [G1397A-T28688C-G29742T] substitutions [4]. These genomes were spread across different geographical regions including; Alborz (n=2), Gilan (n=4), Khuzestan (n=3), Qom (n=1), Sistan and Baluchestan (n=3), Semnan (n=1), South Khorasan (n=1), Tehran (n=18) and unknown (n=3). This indicates that the [B.4/19A] cluster originated very early and began circulating around the country thereafter, reflecting multiple local transmissions. Additionally, there are 2 samples with [B/19A] lineage in this older cluster. These samples were collected back in early March, showing that in the first phase of pandemic; at least two different lineages entered into the country. The [B/19A] samples also carry the G1397A and G29742T substitutions but not T28688C.

The red cluster is comprised of 9 genomes all with the [B.1.*/20A] lineage and collected after May 15^th^, compatible with the pattern shown in Figure 1. The [B.1.*/20A] samples did not show [G1397A-T28688C-G29742T] substitutions but instead harbored [C241T-C3037T-C14408T-A23403G] or [C241T-C3037T-C14408T-A23403G-G25563T], which are the common patterns of variant co-occurrence of B.1 and B.1.* lineages in Europe and North America [21].

#### TMRCA Estimates

The median date of the time to the most recent common ancestor (TMRCA) of 45 high-quality SARS-CoV2 genomes from Iran was estimated as 2019-12-26 with 95% highest posterior density (HPD) intervals of [2019-11-17 to 2020-01-16].

Additionally, to track the appearance of [B.1.*/20A] cluster in the country, the TMRCA of the nine B.1 samples, placing other samples as outgroup was estimated. The TMRCA was 2020-02-22, with HPD intervals of [2020-01-12 to 2020-03-29]. Clearly, more high-quality B.1* genomes are required to narrow the credible interval and predict a more precise TMRCA for entry of this lineage. However, the above results still indicate that the new lineage might have been introduced separately, after the entry of B.4, and then gradually increased in the population, becoming detectable in our cohort since mid-May.

#### Sources of SARS-CoV-2 entries

Subsequent analysis of SARS-CoV-2 genomes from the Iranian outbreak in the context of 216 set of genomes from around the world, clarified the location of the two main clusters among the global samples (Figure 3).

**Figure3.**
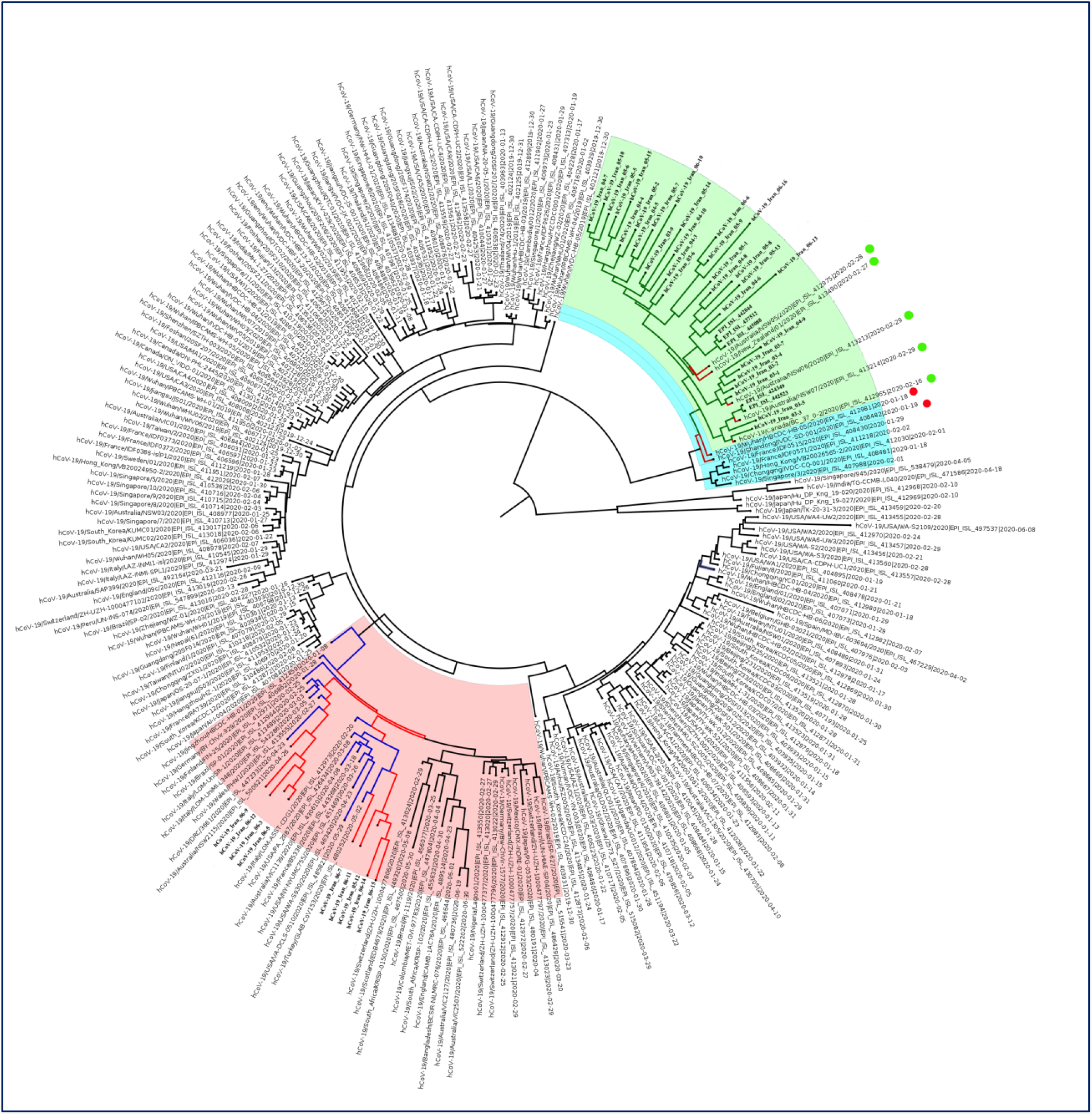
Radial phylogenetic tree of SARS-CoV-2 genomes from the Iranian outbreak in the context of 216 set of genomes from around the world. The major B.4 cluster in the Iranian SARS-CoV-2 outbreak is highlighted in light green. The [B.1.* / 20A] cluster is highlighted in light red. The red circles denote the two B.4 samples from China located near the major Iranian cluster. The green circles denote the five B.4 samples from Australia, Canada and New Zealand located within the major Iranian cluster.

As expected, the B.4 cluster (highlighted in light green) was linked to the early samples collected in January-February and mostly in China. This supports the hypothesis of very early virus introduction to Iran and most likely from China, which is consistent with Iran’s health ministry statements that the virus was brought from China by travelers[22].

Interestingly, the Iranian B.4 cluster is closely linked to the two B.4 samples collected on mid-January (2020-01-19 and 2020-01-18) in Hubei/Wuhan and Shandong/Qingdao in China (EPI_ISL_408482 and EPI_ISL_412981). This suggests that the three [G1397A-T28688C-G29742T] substitutions occurred before their introduction to Iran, subsequently becoming the major lineage and driving the epidemic in the country (Figure 3, Figure 4).

**Figure4.**
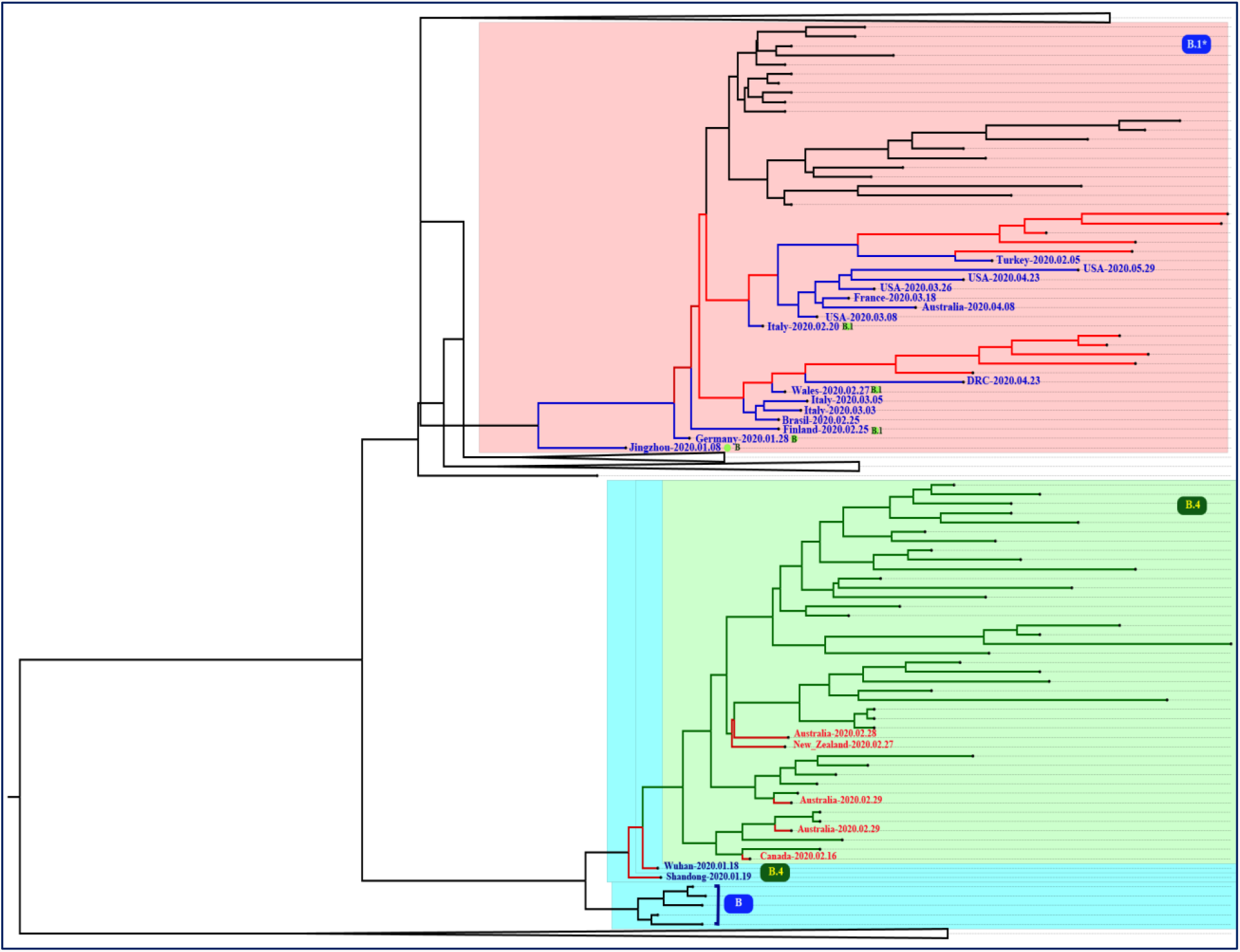
The zoomed and collapsed phylogenetic tree of SARS-CoV-2 genomes from the Iranian outbreak in the context of 216 set of genomes from around the world. The two main clusters circulating in Iran are zoomed. The major B.4 cluster in SARS-CoV-2 Iranian outbreak is highlighted in light green and the lines corresponding to Iranian samples within this cluster are also shown in green while other global samples are shown in red lines. The [B.1.* / 20A] cluster is highlighted in light red and the lines corresponding to Iranian samples within this cluster are also shown in red while other global samples are shown in blue lines.

Afterwards, the virus was transferred to the other countries, such as Canada, Australia, and New Zealand, by travelers [4]. This is now confirmed by locating five samples from these countries within the major Iranian cluster (EPI_ISL_412965, EPI_ISL_413213, EPI_ISL_412975, EPI_ISL_413214, and EPI_ISL_413490). All these five samples were collected in late February, having a travel history to Iran (Figure 3, Figure 4).

The [B.1.*/20A] cluster (highlighted in light red) localizes in a completely different position, namely among the samples from various parts of the world, prominently Europe (Figure 3, Figure 4). This corroborates the hypothesis of new sources of virus introduction to the country (probably in late February as suggested by TMRCA of B.1* samples) before suspension of air flights; as foreign and Iranian airlines transferring passengers between Iran, Europe and North America were suspended on February 25^th^ and March 8^th^, respectively.

### Variant analysis of SARS-CoV-2 genomes of Iranian outbreak

#### Common SARS-CoV2 variants

We detected 14 different variants with >10% frequency in SARS-CoV-2 genomes from Iranian outbreak (Table1). Notably, just four variants, namely G1397A, T28688C, G29742T and G11083T, contributed to >70% of samples; comprising the common co-occurrence of variants; [G1397A-T28688C-G29742T] in B.4 lineage.

**Table 1.** Common and novel variants observed in SARS-CoV-2 genomes of the Iranian outbreak

| No. | Genomic change | Type of mutation | Gene/protein | Amino acid change | No. of samples | Sample Lineages | Description |
| --- | --- | --- | --- | --- | --- | --- | --- |
| Common SARS-CoV-2 variants in Iranian outbreak |  |  |  |  |  |  |  |
| 1 | G1397A | Nonsynonymous | nsp2 | V198I | 43 (81%) | B.4, B | Known coexistence of variants, constituting B.4 lineage, as suggested by Eden et al. 2020. |
| 2 | T28688C | Synonymous | N | L139L | 41 (77%) | B.4 |  |
| 3 | G29742T | Non-coding | 3'UTR | NA | 43 (81%) | B.4, B |  |
| 4 | G11083T | Nonsynonymous | nsp6 | L37F | 39 (74%) | B.4, B | The most common mutation in Asia, during December 2019-March 2020. |
| 5 | C241T | Non-coding | 5'-UTR | NA | 10 (19%) | B.1* | Known coexistence of variants, constituting B.1 (G) and B.1.* (GH) clades, according to Mercatelli and Giorgi 2020. |
| 6 | C3037T | Synonymous | nsp3 | F106F | 10 (19%) | B.1* |  |
| 7 | C14408T | Nonsynonymous | nsp12 (RdRp) | P323L | 10 (19%) | B.1* |  |
| 8 | A23403G | Nonsynonymous | S | D614G | 13 (24.5%) | B.1* (n=10), B.4 (n=3) |  |
| 9 | G25563T | Nonsynonymous | ORF3a | Q57H | 10 (19%) | B.1*, B.4, B |  |
| Unique SARS-CoV-2 haplotypes in Iranian outbreak |  |  |  |  |  |  |  |
| 10 | G20887A | Nonsynonymous | nsp16 | G77R | 9 (17%) | B.4 | New coexistence of variants, observed in same nine samples also carrying B.4 common variants. |
| 11 | C28830T | Nonsynonymous | N | S186F | 9 (17%) | B.4 |  |
| 12 | C21627T | Nonsynonymous | S | T22I | 9 (17%) | B.4 |  |
| 13 | G8653T | Nonsynonymous | nsp4 | M33I | 6 (11%) | B.4 | New coexistence of variants, observed in same six samples also carrying B.4 common variants. |
| 14 | C884T | Nonsynonymous | nsp2 | R27C | 6 (11%) | B.4 |  |
| Unique SARS-CoV-2 variants in Iranian outbreak |  |  |  |  |  |  |  |
| 15 | C28388G | Nonsynonymous | N | Q39E | 1 | B.1.1/20B | Known variants located at the same position: Q39L/ Q39H/ Q39R/ Q39* |
| 16 | G18712A | Nonsynonymous | nsp14 | A225T | 1 | B.4/19A | Known variants located at the same position: A225D / A225S |
| 17 | T3926C | Nonsynonymous | nsp3 | S403P | 1 | B.4/19A | Known variants located at the same position: S403L / S403A |
| 18 | G6461A | Nonsynonymous | nsp3 | V1248M | 1 | B.4/19A | Known variants located at the same position: V1248G / V1248L |

**N:** Nucleocapsid phosphoprotein, **nsp2:** Non-Structural protein 2, **nsp3:** Predicted phosphoesterase, papain-like proteinase, **nsp4:** Transmembrane protein, **nsp6:** Transmembrane protein, **nsp14:** 3'-to-5' exonuclease, **nsp16:** 2'-O-ribose methyltransferase, **ORF:** open reading frame, **RdRp:** RNA-dependent RNA polymerase, **S:** Spike glycoprotein

The G11083T variant is one of the most frequent mutations observed in Asia from December 2019 till March 2020 [21, 23] and, not surprisingly, is observed mostly along with the 3 substitutions constituting the B.4 lineage.

Other less frequent variants were observed in ∼19% of samples constituting the known co-occurrence of variants; [C241T-C3037T-C14408T-A23403G] and [C241T-C3037T-C14408T-A23403G-G25563T], occurring in clade G (B.1) which is prevalent in Europe, Oceania, South America, and Africa and clade GH (B.1.*), prevalent in North America [21].

#### Novel SARS-CoV-2 variants

Remarkably, we observed two specific haplotypes; the co-occurrence of [G20887A-C28830T-C21627T] and [G8653T-C884T] variants in 17% and 11% of samples, respectively (Table1). Both groups also carried B.4 [G1397A-T28688C-G29742T] variants. These haplotypes are less frequent in CoV-GLU and 48,635 genomes investigated by Mercatelli & Giorgi [10, 21]. Therefore, we conclude that the observation of these variants with the [G1397A, T28688C, and G29742T] is not a common feature worldwide. However, further investigations are required to assess their extent of significance into SARS-CoV2 genetic diversity in Iran.

Analysis of viral isolates also revealed 4 samples harboring unique variants (Table1), not detected in sequences from SARS-CoV-2 pandemic in CoV-GLUE and also not in 48,635 complete genomes investigated by Mercatelli and Giorgi [10, 21]. Among these, the C28388G (Q39E) variant is located in Nucleocapsid phosphoprotein (N), which is one of the essential SARS-CoV2 proteins having role in packaging the RNA genome of the virus into capsid and therefore playing an important role in SARS-CoV2 self-assembly [24].

Although the exact variants were unique, different missense variants at the same location were identified in other viral sequences around the world.

#### Variants located in Spike (S) protein

Spike glycoprotein is the key protein mediating the entry of the virus to the cell and therefore the target of most vaccine strategies [25]. We thus focused on variants located in the spike protein of viral isolates from Iran. We identified spike variants in 28 samples (53%), in which D614G and T22I variants were occurring at higher frequencies of 24.5% and 17%, respectively (Table2).

**Table 2.**
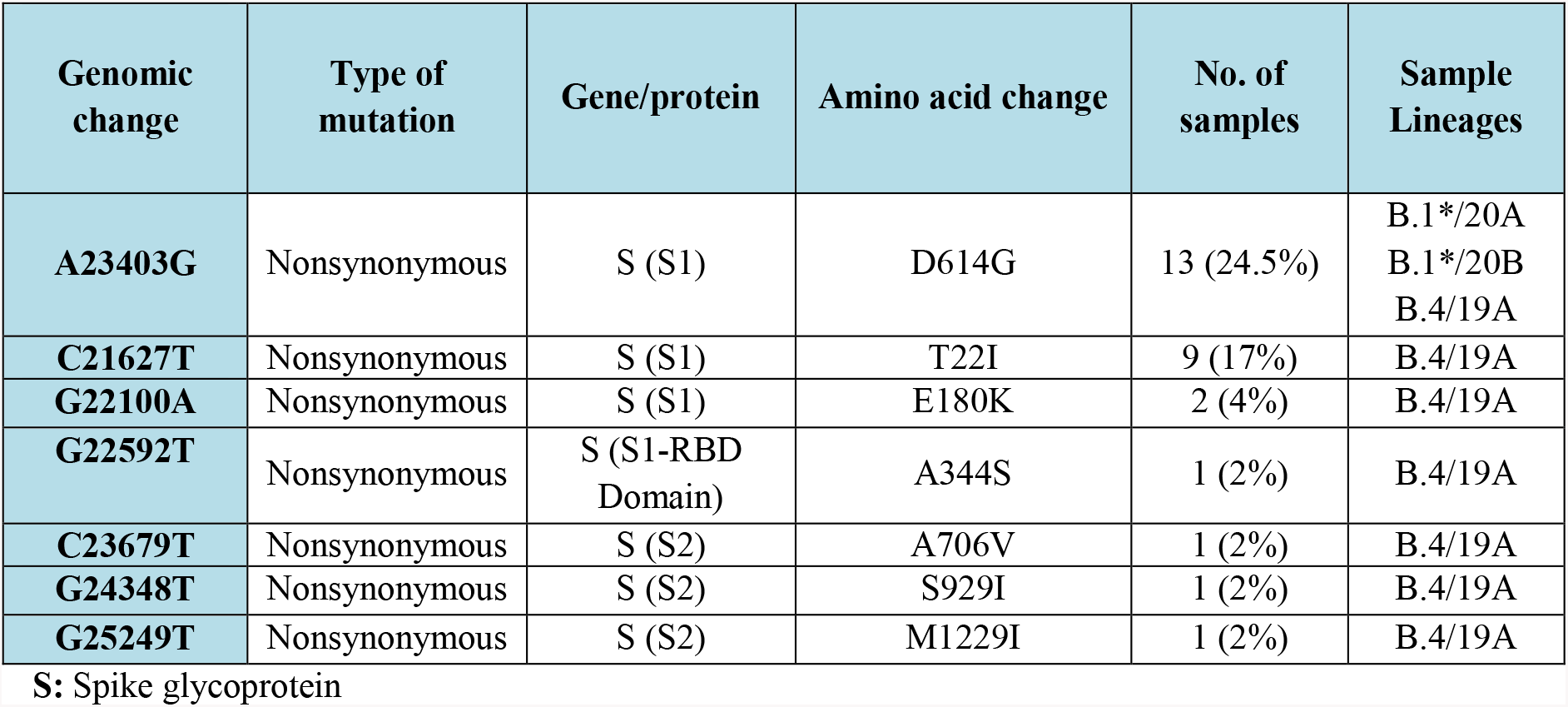
Variants located in the spike, observed in SARS-CoV-2 genomes of the Iranian outbreak

| Genomic change | Type of mutation | Gene/protein | Amino acid change | No. of samples | Sample Lineages |
| --- | --- | --- | --- | --- | --- |
| **A23403G** | Nonsynonymous | S (S1) | D614G | 13 (24.5%) | B.1*/20A<br>B.1*/20B<br>B.4/19A |
| **C21627T** | Nonsynonymous | S (S1) | T22I | 9 (17%) | B.4/19A |
| **G22100A** | Nonsynonymous | S (S1) | E180K | 2 (4%) | B.4/19A |
| **G22592T** | Nonsynonymous | S (S1-RBD Domain) | A344S | 1 (2%) | B.4/19A |
| **C23679T** | Nonsynonymous | S (S2) | A706V | 1 (2%) | B.4/19A |
| **G24348T** | Nonsynonymous | S (S2) | S929I | 1 (2%) | B.4/19A |
| **G25249T** | Nonsynonymous | S (S2) | M1229I | 1 (2%) | B.4/19A |

S: Spike glycoprotein

The T22I was only observed in B.4/19A cluster and the D614G mutation occurred mostly in [B.1.* / 20A] cluster.

The D614G variant, being the most prevalent mutation in the sequenced genomes around the world, was also the most prevalent spike mutation in viral isolates from Iran, while showing an increasing trend from mid-May and locating mostly within the [B.1.* / 20A] cluster. In accordance with this, most of our samples with D614G mutation were harboring [C241T-C3037T-C14408T] or [C241T-C3037T-C14408T-G25563T].

Remarkably, co-occurrence of D614G mutation with B.4 [G1397A-T28688C-G29742T] and G11083T variants was also observed in three samples.

Moreover, Sanger sequencing of additional 67 SARS-CoV2 positive samples confirmed the increase in D614G frequency till October, becoming the dominant mutation in the Iranian outbreak (Figure 5).

**Figure5.**
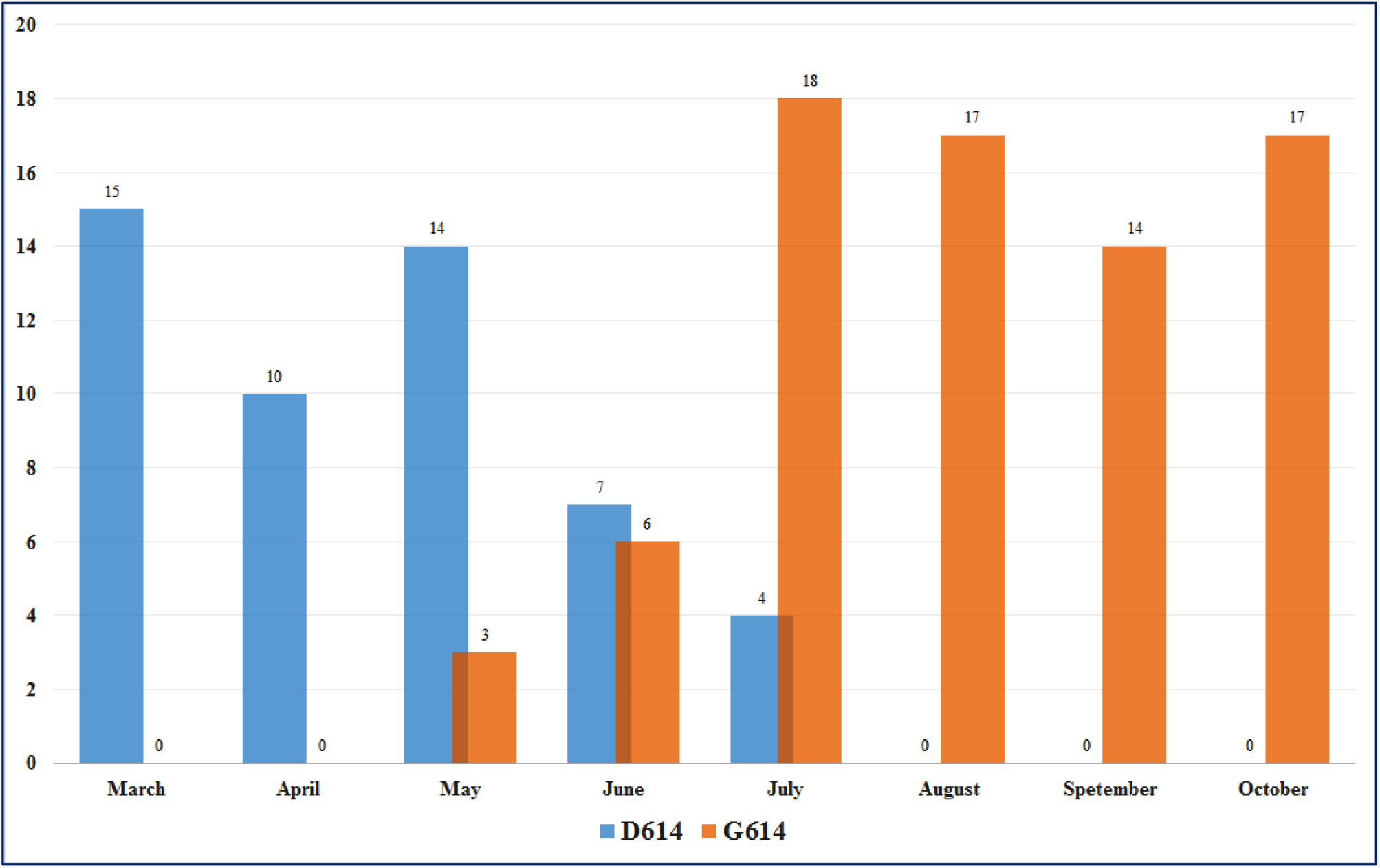
The frequency of D614G mutation during March-October interval in Iranian SARS-CoV-2 outbreak

## Discussion

The current study is the first comprehensive analysis of SARS-CoV-2 full genomes in Iranian outbreak. Regarding the importance of real-time sequencing of emerging viruses, and lack of full genome in Iran, this study was designed to provide 50 SARS-CoV-2 full genome sequences of the epidemic in Iran.

Lineage assignment and phylogenetic analysis of these sequences clarified the origins and transmission dynamics of SARS-CoV-2 outbreak in Iran, in which two major introductions into the country was detected, constructing two major clusters of the virus in different times, followed by rapid community transmission throughout the country.

This study confirms the B.4 as the dominant lineage in Iran, from the start of the epidemic till the end of June. This lineage was primarily introduced as a prevalent distinct clade in Australian travelers from Iran in early April [4]. Since then, it is recognized as the Iranian epidemic. Nonetheless, no specific study on Iranian SARS-CoV-2 samples was performed so far, which is addressed in this study.

Furthermore, the estimated median date of the time to the most recent common ancestor in this study indicates the possibility of cryptic transmission of the virus or presence of misdiagnosed patients for more than one month prior to the report of first cases of the COVID-19 disease in the country.

Therefore, the B.4 lineage originated first in late-2019/early-2020 followed by multiple local transmissions, developing the major SARS-CoV-2 clade in the beginning of outbreak. Our data suggest a reduction in B.4 dominancy, followed by a surge of B.1.* lineages which has been exported from Europe globally. This new cluster might be explained by new sources of virus entries before suspension of flights in late February or being the mutant product of B lineage in early phase.

In addition to outbreak tracing, these full genome sequences can provide beneficial information in understanding the genome diversity of viral isolates in Iran, helpful in adapting more specific diagnostic tests, therapeutic approaches and vaccines.

Generally, RNA viruses are known to have a high mutation rate, which is explained by lack of proofreading activity of RNA-dependent RNA polymerase (RdRP). However, coronaviruses are among the exceptions, with lower mutation rates due to the presence of RdRp-independent proofreading activity [26, 27]. The mutation rate of 1.12 × 10^−3^ mutations per site-year [23] and the average number of 7.23 mutations per sample [21], are in support of the moderate mutation rate of SARS-CoV-2, similar to SARS-CoV-1.

Variation tracking of the viral isolates has shown that some SARS-CoV-2 mutations-such as P323L and D614G-are distributed globally while some other ones are accumulated in specific geographical regions [28].

Investigation of prevalent mutations in Iran indicated the co-occurrence of some widespread mutations consistent with the two main lineages in the country. Furthermore, the unique mutations and also haplotypes of [G20887A-C28830T-C21627T] or [G8653T-C884T] along with the known [G1397A-T28688C-G29742T] mutations, were identified in low proportion of samples. Therefore, none of those could be considered as adapted geographically in Iranian SARS-CoV-2 samples. Indeed, massive sequencing of a larger cohort is a requisite for investigating the significance of these variants. Moreover, the impact of these country-specific variants (yet to be defined) on the behavior of virus (replication efficiency, virulence. etc.) needs further investigation.

Despite the relatively low mutation rate of SARS-CoV-2 genome, still a total of 353,341 mutations were identified in 48,635 SARS-CoV-2 genomes compared to the NC_045512.2 as of July 2020 [21]. Among these, studying the mutations in the spike protein are of great importance, as this immunogenic structural protein mediates the virus entry to the host cells via interacting with cellular receptors such as angiotensin-converting enzyme 2 (ACE2) and as it plays a key role in induction of neutralizing antibodies [29, 30]. This glycoprotein is composed of two functional subunits, S1 and S2. The S1 subunits contain a receptor binding domain (RBD) through which SARS-Co-2 binds to the ACE2 receptor, while S2 is responsible for virus-host membrane fusion [31]. Furthermore, the spike protein is the target for many vaccine candidates currently in development and therefore tracking the mutations in this protein (especially RBD region) is crucial [30].

Monitoring the spike mutations in Iranian SARS-CoV-2 genomes revealed no novel mutations in this genomic region, while determined two commonly known mutations; T22I and D614G. Both of these variants are outside RBD region, suggesting no negative effect for efficacy of the future vaccines on the viral lineages currently circulating in Iran, similar to other countries as discussed by Dearlove et al. [30].

While the known T22I mutation is less frequent in other regions, the D614G variant is now the most prevalent mutation in COVID-19 pandemic [21, 25, 27, 32, 33]. Recent studies suggested a fitness advantage for G614, rapidly making it the dominant form in each geographical location [25, 32]. This might be the result of a significant conformational change in spike protein caused by G614, leading to more feasible virus-host cell membrane fusion. As a consequence, increased infectivity, transmission and replication fitness is reported for G614 [33]. However, there are still some debates about the transmission effect of D614G variant [34, 35] but it is recently shown that the mutation is not related to increased disease severity and does not reduce the effect of neutralizing antibodies [32, 36].

Notably, in accordance with the increasing [B.1*/20A] clade among the viral isolates of the Iranian epidemic as of mid-May, the D614G variant is dominating in the population. This can partly explain the accelerated transmission of the virus in recent months, although the negative effect of relaxed quarantine policies should not be overlooked.

Due to the strong fitness of G614, we could also observe its co-occurrence with common B.4 variants, while the mutation is known as always co-occurring with variants defining the G clade [21].

In conclusion, genomic sequencing and phylogenetic analysis suggested that SARS-CoV-2 entered to Iran, in very late 2019 and circulated among vulnerable patients. Now, the increase in frequency of D614G mutation during the course of pandemic predicts a critical situation and raises a significant warning alarm for considering stronger prohibition strategies in the country.

## Data Availability

Some of the sequence data is being uploaded in GISAID database and the rest is in process within 2 days.

https://www.gisaid.org/

## Acknowledgments

Iranian Network for Research in Viral Diseases (INRVD) is acknowledged for help with the sample and data collection through its collaborative research centers and hospitals. This study was funded by Iran Vice deputy for Research and Technology at Iran Ministry of Health and Medical Education, grant number: 99/801/A/6/7586.

